# Detection of Hemagglutinin H5 influenza A virus RNA and model of potential inputs in an urban California sewershed

**DOI:** 10.1101/2024.12.31.24319823

**Authors:** Abigail P. Paulos, Stephen P. Hilton, Bridgette Shelden, Dorothea Duong, Alexandria B. Boehm, Marlene K. Wolfe

## Abstract

In 2024, the highly pathogenic avian influenza A H5N1 caused outbreaks in wild birds, poultry, cows, and other mammals in the United States with 61 human cases also reported by the CDC. Detection of influenza A H5 RNA in wastewater has been previously reported in sewersheds in Texas and North Carolina with nearby impacted dairy herds following the emergence of H5N1 in dairy cows. Here, we conduct retrospective testing of total influenza A and H5 hemagglutinin genes in wastewater as well presenting and applying new assays for detection of H1 and H3 genes across a respiratory virus season in an urban California sewershed from September 2023 – May 2024. Total influenza A, H1, and H3 were regularly detected, while H5 was first detected in March. We developed a model that uses Monte Carlo simulations and previously published parameters to estimate numbers of infected people, poultry, wild birds, or liters of H5-contaminated milk required to result in measured H5 concentrations in wastewater. Our findings demonstrate that in this California sewershed, contaminated milk or infected poultry were the most likely sources of H5 to wastewater. We created a publicly available tool to apply the H5 input model in other sewersheds estimate required inputs.

**Synopsis:** We developed a model to understand potential sources of influenza A H5 RNA in wastewater, enabling interpretation of H5 RNA wastewater detections.

*TOC figure:* 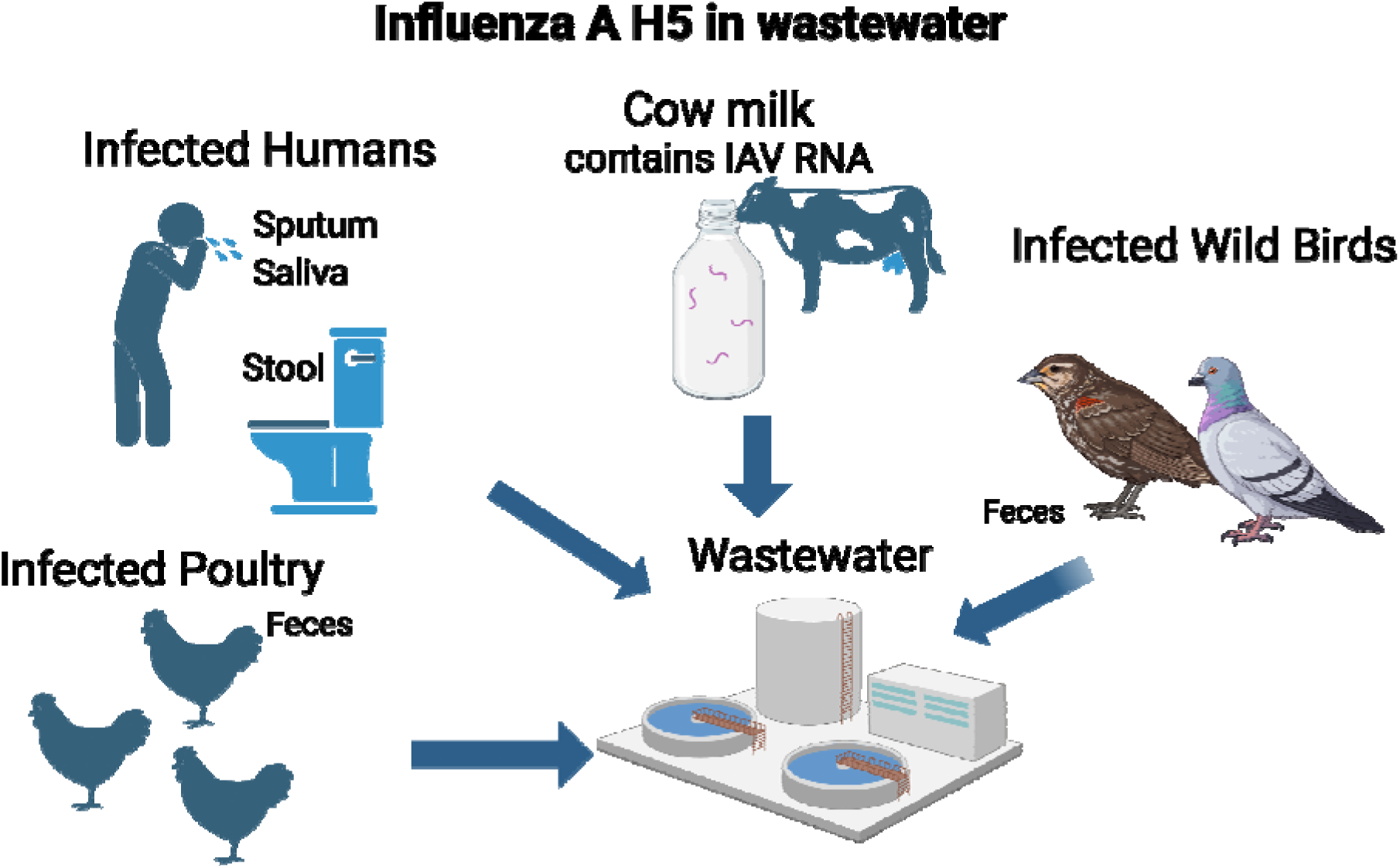

## Introduction

Approximately 3-11% of the population in the United States is infected with influenza virus each year, and Influenza A virus (IAV) is responsible for the majority of these infections.^1^ In recent years, wastewater monitoring of infectious diseases has emerged as a powerful tool for tracking trends in disease incidence in the community. Previous research has found that concentrations of total IAV genomic RNA in wastewater track closely with occurrence of infections in the contributing community.^2–5^ IAV has subtypes that are classified by both hemagglutinin (H) and neuraminidase (N) proteins.^6^ The H1 and H3 subtypes are most common in humans and swine, while many subtypes, including H1, H3, H5, H7, H9, and H10, are common in avian populations.^7^ Given that levels of total IAV (measurements that include all H subtypes) in wastewater have been closely correlated with human disease, and municipal wastewater primarily consists of anthropogenic inputs, the predominant source of IAV RNA in wastewater has thus far been considered to be human.

The highly pathogenic avian influenza (HPAI) A (H5N1) outbreak in birds began in the United States in February 2022 and has resulted in the deaths of millions of poultry and wild fowls.^8^ This outbreak in poultry has prompted concerns about increased spillover into humans and the possibility of future human to human transmission, as humans sporadically infected with H5N1 have experienced severe symptoms and high mortality in the past.^9^ Other animal influenza strains have jumped from animal populations to humans through mutations, adaptation, or reassortment events with human influenzas, resulting in epidemics and pandemics.^10,11^ This concern has grown as outbreaks of unknown illness in cattle in March 2024 were identified as infections caused by H5N1 later that month. Between March 1 and December 19, 2024, 865 infected dairy cow herds and 61 human cases were reported in 16 states.^12^ Human cases of H5N1 were reported mostly among those who worked in close proximity to dairy cows and poultry; human-to-human transmission has not been reported or suspected in any of the known cases.^13–15^

Beginning in March 2024 and concurrent with cattle outbreaks, we detected the H5 IAV subtype genomic RNA (hereafter, H5 RNA) in wastewater in multiple sites across the United States.^16^ Prior to the H5N1 outbreaks in cattle, contributions to wastewater from animals infected with H5N1 (such as poultry) were expected to be minimal. However, cows infected with H5N1 shed the virus in milk, and IAV RNA has been detected both in raw milk and in pasteurized milk available on store shelves.^17,18^ Because of the food chain, a substantial amount of milk products enter sewers; it is estimated that 12% of milk available for sale is wasted at retailers and 20% by consumers after purchase.^19^ Milk is commonly provided with breakfast and lunch to students at schools, and much of this milk is wasted as well. Studies report that 45% of milk was wasted at kindergarten and pre-kindergarten lunches,^20^ and 13% by high school students.^21^ This wasted milk may be disposed of down the drain, entering sewer systems and contributing detectable viral RNA to wastewater. Food industries are also often permitted to dispose of waste generated while processing dairy products into sewer systems. Contributions to wastewater from birds are less likely because of the lack of connection between wild bird habitats and domestic flocks to municipal sewer systems. However, wastewater sewer systems that accept stormflow (“combined sewers”) are less common but may receive fecal waste from wildfowl and other wild birds that mixes with runoff and enters sewer systems. Both poultry and wildfowl can shed IAV in their feces when infected.^22,23^ Wastewater treatment plants with open-air settling tanks, or that accept waste from industries that generate standing water, may be susceptible to inputs of bird feces.

Therefore, we conclude that any one of cow’s milk, poultry feces, wildfowl feces, or human contributions are the most likely sources of H5 in wastewater.

In this study, we present findings from retrospective testing of wastewater solids for total IAV and H1, H3, and H5 IAV subtype RNA from September 2023 - May 2024 in a publicly owned treatment work (POTW) in California. We also present findings from a model developed to estimate the theoretical contributions of humans, cow milk, wildfowl feces, and poultry feces to the wastewater system necessary to result in the observed H5 RNA concentrations. Our objectives were to 1) identify the timing of the first H5 detection in the sewershed during a period with circulating human influenza and 2) develop and implement a model of theoretical sources of H5 into the wastewater system to contextualize our findings.

## Methods

### H1 and H3 Assay Design

IAV H1 and H3 subtype genome sequences were downloaded from the National Center for Biotechnology Information (NCBI) in January 2023 (H1 genomes) and December 2022 (H3 genomes) and supplemented with additional newer sequences available from NCBI and GISAID in April 2024. Sequences were aligned, and primers and probes were designed to target the hemagglutinin (HA) gene using Primer3Plus. Parameters used in assay development (e.g., sequence length and GC content) are provided elsewhere.^24^ The primers and probes (Table S1) were confirmed to be specific and sensitive for influenza A containing the H1 and H3 subtypes of the HA gene *in silico* using NCBI BLAST.

The H1 and H3 assays were tested *in vitro* against nucleic acids from a large collection of respiratory pathogens including different influenza subtypes (Table S2). Nucleic acids from these panels were extracted and purified as described below for wastewater solids samples, then used as template in droplet digital RT-PCR assays. The panels were run in a single well using the same ddPCR methods described below and elsewhere.^16,24^

### IAV M gene and H5 assays

The H5 assay used has been described in detail elsewhere,^16^ as has the M gene assay.^24^ See the Supporting Information for information on primers, probes, and positive control material. The influenza M gene assay detects all IAV subtypes and hereafter, measurements made using the M gene assay will be interpreted as “total IAV RNA”.

### Retrospective analysis of samples

Biobanked nucleic-acid extracts obtained from wastewater solids samples collected between September 1, 2023 and May 13, 2024 from a POTW (Southeast San Francisco) were retrospectively analyzed for total IAV (M gene), H1, H3, and H5 (n=110 samples). Nucleic-acids were stored between 1 and 10 months at -80°C before analysis. Samples consisted of grab samples of settled solids from the primary clarifier. Samples were collected using sterile methods and stored at 4°C prior to nucleic-acid extraction. The POTW is located in an urban area and serves 750,000 people, and services a geographic area with combined stormwater and sanitary sewer systems meaning that stormwater and urban dry weather runoff (generated via irrigation or car washing, for example) may enter the POTW.

Sample processing methods are described in detail elsewhere.^24–26^ Briefly, nucleic acids were extracted from the solid fraction of each sample using the Chemagic Viral DNA/RNA 300 Kit H96 (PerkinElmer, Shelton, CT) followed by inhibition removal (Zymo OneStep PCR Inhibitor Removal Kit, Irvine, CA).^25^ Nucleic-acid extraction occurred immediately within 24 h of sample collection. Between 0.5-1g of the dewatered solids were dried at 110°C for 19-24 h to determine the dry weight. Concentrations of total IAV (M gene), H1, H3, and H5 RNA were measured in multiplex using droplet digital 1-step RT-PCR (dd-RT-PCR) run on an AutoDG Automated Droplet Generator (Bio-Rad, Hercules, CA), Mastercycler Pro (Eppendorf, Enfield, CT) thermocycler, and a QX600 Droplet Reader (Bio-Rad). Ten ddPCR replicates were run for each sample. Only wells with over 10,000 droplets were included; all wells met this criterion. Extraction and PCR positive and negative controls were run on each 96-well plate. Positive controls consisted of Twist synthetic controls (M gene, H1, H3) and custom gene blocks (H5; IDT); see the SI for full details. Negative controls consisted of no template controls (NTC) containing nuclease free water. Bovine coronavirus (BCoV) was spiked into samples and measured as an internal control, and pepper mild mottle virus (PMMoV) was measured using ddPCR as an endogenous internal control. We used the QuantaSoft Analysis Pro software (Bio-Rad) to threshold droplets, and a sample had to have 3 or more positive droplets to be considered positive. Three positive droplets corresponds to a concentration between 500-1000 copies/g. Values were converted to gene copies/dry g using dimensional analysis. See the SI for reaction chemistry and cycling parameters.

### Model of theoretical inputs

We developed a model to estimate the H5 RNA inputs into the system that would be required to result in the measured H5 RNA concentrations in wastewater solids. We considered inputs from humans, cow milk, poultry feces, and waterfowl feces, as each input type had a plausible mechanism for entering wastewater. We note that these inputs may enter wastewater in a number of ways, including from household uses, businesses, industrial discharges, and, in some cases, environmental sources.

**Figure 1.**
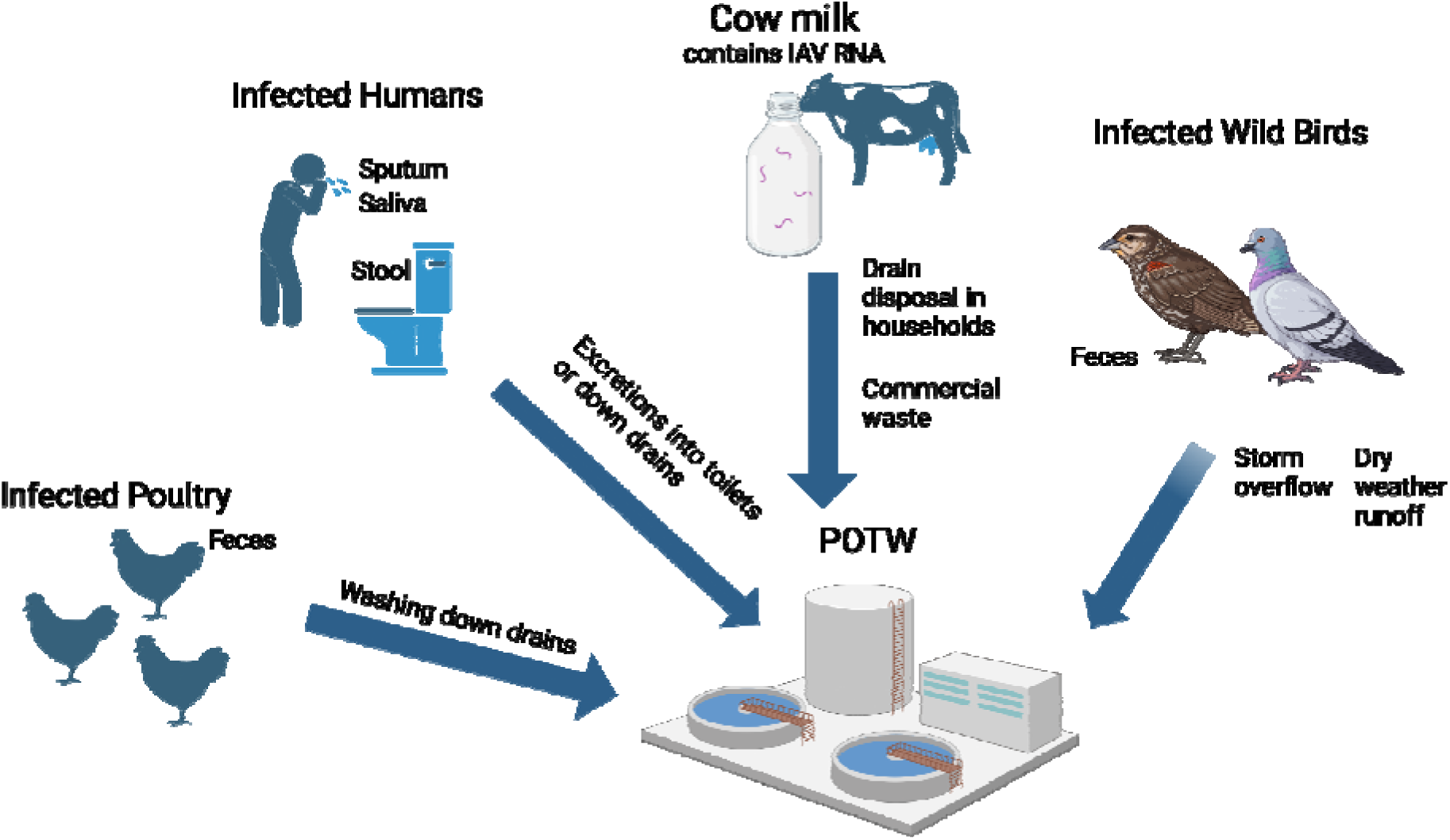
Conceptual image of sources of H5N1 to the POTW.

### Mass Balance

We assume that the concentrations measured in wastewater relate to inputs from humans, birds, and cow’s milk according to the following equation:

where indicates the daily flux of H5 RNA (gene copies/day) through the POTW and , , , and indicate the daily contribution (gene copies/day) of humans, poultry, wild birds, and milk, respectively, to the POTW.

*Calculations of daily H5 wastewater fluxes,*

We assume that both the liquid and solid portions of wastewater contain H5 RNA when detected in solids and calculate total H5 RNA content across both phases. The concentration in liquids is calculated using the Freundlich isotherm:^27^ where indicates the H5 RNA concentration in liquids (gene copies/mL), indicates the H5 RNA concentration in solids (gene copies/dry gram), and indicates the Freundlich partitioning coefficient (mL/dry gram).

Total daily H5 RNA flux through the POTW (gene copies/day) is calculated as the sum of H5 RNA in solids (gene copies/day) and in liquids (gene copies/day). We obtain the daily flow rate (in millions of gallons per day) and total suspended solids (TSS; mg/L) from the POTW for each day. The total daily flux of solids through the POTW is calculated as:

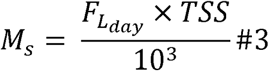

where F_L_day_ is the daily flow rate in terms of L/day, TSS is the total suspended solids (mg/L), and the 10^3^ factor is to convert from mg/day to g/day. The daily flux of H5 RNA in the solids (gene copies/day) is calculated by:

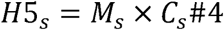

The daily flux of H5 RNA in the liquids (gene copies/day) is calculated by:

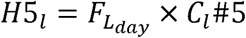

The total daily flux of H5 RNA through the POTW (gene copies/day) is calculated by:

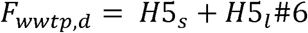

### Human inputs

We consider human contributions to the POTW from saliva, sputum, and feces, such that:

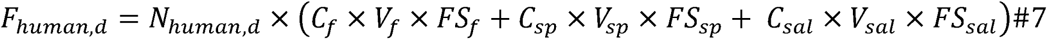

where *N* indicates the number of infected people contributing to wastewater, *C* indicates concentration in that excretion type (gene copies per mL or per g), *V* indicates volume of that excretion type entering the wastewater system per day per person (mL or g), and *FS* indicates the fraction of infected persons shedding influenza by that route (unitless). *F* refers to feces, *sp* refers to sputum, and *sal* refers to saliva.

Assuming no contribution to the POTW from birds or milk, we can simplify Eq. 1 to:

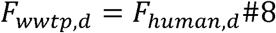

and, plugging in Eq. 7:

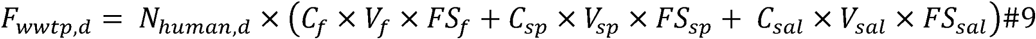

Rearranging to solve for the number of infected individuals (*N*):

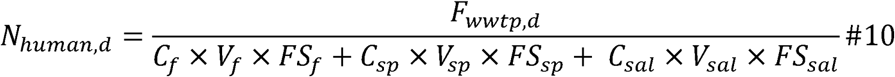

### Poultry inputs

We consider inputs from poultry to the POTW through feces:

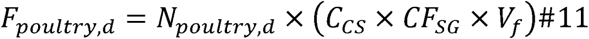

Where c_CS_ indicates the concentration of influenza reported in cloacal swabs collected from domestic poultry (gene copies/swab), cF_SG_ indicates a conversion factor from concentration/swab to concentration/g (swab/gram), and v_f_ indicates the volume of feces produced by birds per day (grams).

Assuming no contribution to the POTW from humans or milk, we can simplify Eq. 1 to:

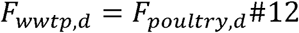

and, plugging in Eq. 11:

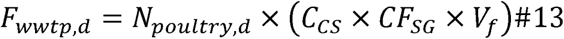

Rearranging to solve for the number of infected birds (*N*):

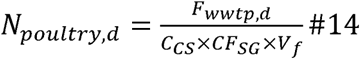

### Wild bird inputs

We consider inputs from wild birds to the POTW through feces:

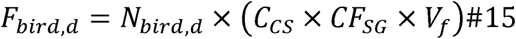

Where c_CS_ indicates the concentration of influenza reported in cloacal swabs collected from domestic poultry (gene copies/swab), cF_SG_ indicates a conversion factor from concentration/swab to concentration/g (swab/gram), and v_f_ indicates the volume of feces produced by birds per day (grams).

Assuming no contribution to the POTW from humans or milk, we can simplify Eq. 1 to:

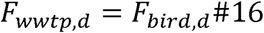

and, plugging in Eq. 15:

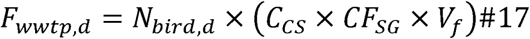

Rearranging to solve for the number of infected birds (*N*):

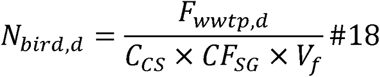

### Cow milk inputs

We estimate the daily flux of H5 RNA from cow milk (gene copies/day) by:

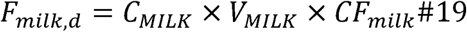

where c_MILK_ refers to the H5 concentration in cow’s milk, v_MILK_ indicates the volume of milk entering the wastewater system per day, and cF_milk_ is a conversion factor from the EID50/mL units c_MILK_ is reported in to gene copies/mL units.

Assuming no contribution to the POTW from humans or birds, we can simplify Eq. 1 to:

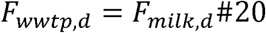

and, plugging in Eq. 19:

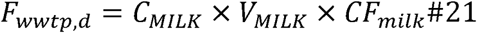

Rearranging to solve for the daily volume of infected milk entering the POTW:

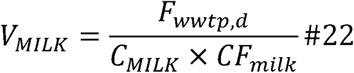

### Data Sources

Using previously published systematic reviews and parameters on daily human waste input into the system, we identified a distribution of human IAV shedding into wastewater per day per infected individual (**Table 1**).^28,29^ For cow milk, we identified a distribution of H5N1 RNA per gallon of pasteurized milk using published reports on H5N1 EID50 content in pasteurized cow milk^17,18^ and a conversion from EID50 to RNA gene copies.^30^ Distributions of daily H5 RNA shedding in the feces of poultry and waterfowl were separately estimated from studies reporting IAV shedding in each bird type (**Table S4**).^22,23^ Blue-winged teal were used as the model species for wild birds, per Humphreys *et al*.^31^ Raw data for empirical distributions are provided in the Stanford Digital Repository (https://purl.stanford.edu/ks454rq5640).

**Table 1.** Model parameters for human wastewater inputs.

| Category | Parameter | Unit | Distribution | Parameters | Reference |
| --- | --- | --- | --- | --- | --- |
| **HUMANS** |  |  |  |  |  |
| *HUMAN WASTEWATER INPUTS* |  |  |  |  |  |
| Sputum | mL sputum produced to sewer per person | mL / day | Uniform | min = 0.1, max = 1 | 29 |
| Saliva | tooth brushing events/day | Tooth brushing events/day | Discrete | [(0,0.02),(1,0.29),(2,0.55),(3+,0.14)] | 29 |
| Saliva | mL saliva to sewer per tooth brushing event | mL/Tooth brushing event | Point | 1 | 29 |
| Stool | stool production per day per person | Log g/day | Truncated Log Normal | meanlog = 4.763, sdlog=0.471, min=0, max=520 | 29 |
| Urine | urine production per day per person | L/day | Gamma | $\alpha$ (shape)=5.315, $\beta$(scale)=0.25, and a (positive) offset $\Delta$ of 0.5 | 29 |
| *HUMAN SHEDDING* |  |  |  |  |  |
| Sputum | gene copies of influenza in sputum | gc / mL | Empirical |  | 28 |
| Saliva | gene copies of influenza in saliva | gc / mL | Empirical |  | assume equivalent to sputum shedding, per findings by Crank *et al* for SARS-CoV-2<sup>29</sup> |
| Stool | gene copies of influenza in stool | gc / g | Empirical |  | 32–35 |
| Urine | gene copies of influenza in urine | gc / mL | Point | 0 | assumption |
| *HUMAN FRACTIONAL SHEDDING* |  |  |  |  |  |
| Sputum | fraction of infected people shedding influenza in sputum | unitless | Uniform | min = 0.73, max = 1 | Calculated from data in Lowry, Wolfe, & Boehm<sup>28</sup> |
| Saliva | fraction of infected people shedding influenza in saliva | unitless | Uniform | min = 0.85, max = 1 | Calculated from data in Lowry *et al.*<sup>28</sup> |
| Stool | fraction of infected people shedding influenza in stool | unitless | Truncated normal | mean = 0.40, sd = 0.21; min = 0, max = 1 | Calculated from data in Lowry *et al.*<sup>28</sup> |
| Urine | fraction of infected people shedding influenza in urine | unitless | Uniform | min = 0.05, max = 1 | Assumption based on Lowry *et al.*<sup>28</sup> |

| **COW MILK** |  |  |  |  |  |
| --- | --- | --- | --- | --- | --- |
| milk | H5 concentration in commercial (pasteurized) milk | log10 EID50 / mL | Truncated lognormal | Mean = 5.93, sd = 2.63, min = 0, max = $2.5 \times 10^5$ | Calculated from <sup>17,18</sup> |
| milk | conversion from EID50 to gc | gc / EID50 | Uniform | Min = 10, max = 25 | <sup>30</sup> |
| milk | conversion from mL to gallons | mL/gallon | Point | 3785.41253 | constant |
| **POULTRY AND WILD BIRDS** |  |  |  |  |  |
| poultry | H5 concentration , cloacal swabs | Log10 gc / swab | Log10 normal | mean = 4.6, sd = 1.6 | Extracted from Fig 3 for chickens<sup>23</sup> |
| waterfowl | H5 concentration , cloacal swabs | Log10 gc / swab | Log10 normal | mean = 3.68, sd = 1.0 | <sup>22</sup> |
| both | mass feces per cloacal swab | g / swab | Uniform | min = 0.05, max = 0.2 | assumption |
| both | conversion factor - swab to g | swab / g | Point | 1 / mass_feces_cloacal_swab | Conversion factor |
| poultry | daily amount of bird poop / bird, g | g / day | Uniform | min = 130, max = 184 | assumption from values in Tanczuk *et al*<sup>36</sup> |
| waterfowl | daily amount<br>of bird poop /<br>bird, g | g / day | Uniform | min = 5.7, max = 8.7 | Calculated<br>from <sup>37</sup> |
| both | fractional<br>shedding | unitless | Point | 1 | assumption |

To estimate the total daily H5 RNA for the POTW, we calculated total solids and total liquids at the POTW per day using the flow rate and total suspended solids (TSS) reported by the POTW. We assumed homogeneity in both solids and liquids, meaning that the quantity of H5 RNA measured in the solids sample is the same throughout all solids passing through the system. The H5 RNA concentration in liquids was estimated from the solids using the partitioning coefficient of IAV RNA in wastewater and the adsorption intensity (**Table 2**).^27^ We summed the total H5 RNA in solids and total H5 RNA in liquids to calculate the total H5 RNA at the POTW per day.

**Table 2.** Wastewater model input parameters.

| Category | Parameter | Unit | Distribution | Parameters | Reference |
| --- | --- | --- | --- | --- | --- |
| wastewater | solid-liquid<br>partitioning<br>coefficient | mL / g | Empirical |  | <sup>27</sup> |
| wastewater | Adsorption<br>intensity | unitless | Uniform | min = 0.7, max = 1.9 | <sup>27</sup> |

Raw data were fit to distributions using the *fitdistr* package in R. For each parameter, we first used the *descdist* function to create a Cullen and Fray graph, which plots the square of skewness against kurtosis. The value from the observed data is plotted, as are the locations of theoretical distributions. Whichever theoretical distribution the observation is plotted nearest is selected as the distribution type, with the beta distribution being the lowest priority (meaning if the observation is near normal, uniform, exponential, or logistic, while still being located within the beta region, the other non-beta distribution is selected). After selecting a distribution type, data were fitted to the distribution using the *fitdist* package and distribution parameters were extracted.

### Model Approach

In the models, we assumed that the measured H5 RNA in wastewater originated entirely from one of the four input types and then independently calculated (1) the number of infected humans in the sewershed, (2) the liters of milk input into the sewer, (3) the number of poultry contributing feces to the sewer, and (4) the number of waterfowl contributing feces to the sewer that would be required to result in the measured H5 concentration. By looking at each source independently, we can estimate an upper bound on the number of infected individuals or IAV-impacted liters of milk that could be contributing to the system. To assess variability in our parameter estimates, we conducted Monte Carlo simulations using n=10,000 iterations. For each iteration, we randomly selected a value from each parameter’s distribution and conducted the calculations. Presented throughout are the median and interquartile range (25th - 75th percentile) from these simulations. Due to the limited data available for our assumptions, we are reporting order of magnitude results.

### Sensitivity Analysis

We use a previously-published method to conduct a sensitivity analysis to determine which parameters the model is most sensitive to.^38,39^ The model is run with the parameter first set to its 25th percentile (p25) then set to the 75th percentile (p75) while all other parameters are set to their median value. We calculate the ratio between the model output for p75/p25. A p75:p25 ratio of 1 indicates that the model is not sensitive to the parameter, while a ratio above or below indicates the model is sensitive to that parameter. For ratios below 1, we calculated the inverse (1/p75:p25) so that all ratios are presented on a comparable scale.

### Shiny Application

We developed an application using the R Shiny package to make the model available for public use. Users can input the H5 wastewater concentration in solids (gene copies/dry g) and the flow rate (in millions of gallons [MGD] per day) and total suspended solids (TSS; mg/L) at the treatment plant. The model will conduct n=50,000 iterations and provide estimates of the required inputs. The Shiny app is available at https://abharv52.shinyapps.io/h5_input_estimates/.

### Identifying food and/or agricultural industries within the sewershed

We used multiple online sources to search for potential industrial contributors of dairy products to the sewershed wastewater. We searched google maps for dairy, cheese, or butter processing facilities and cross-checked locations with the sewershed boundaries to see if they fell within the sewershed. The boundary shapefile was provided by the POTW. We also searched the Enforcement and Compliance History Online (ECHO) database for facilities with Clean Water Act permits with a Standardized Industry Code (SIC) indicating dairy processing (**Table S5**).

## Results

### H1 and H3 assay performance and QA/QC

Both *in silico* and *in vitro* testing indicated the H1 and H3 assays are 100% specific. *In vitro* tests returned non-detects for all non-target pathogens, including other influenza subtypes.

The Environmental Microbiology Minimal Information (EMMI)^40^ and McClary-Gutierrez et al. guidelines^41^ were used for reporting results (both checklists available at https://purl.stanford.edu/ks454rq5640). Positive and negative controls performed as expected. Nucleic acid extraction efficiency in samples exceeded the quality control threshold of 0.1 for bovine coronavirus (median = 1.8, interquartile range (IQR) = 1.2 -2.4); recoveries greater than one are likely a result of errors associated with the measurement of bovine coronavirus seeded into the sample. PMMoV was detected at high concentrations in all samples (median of 6.1 × 10^8^, IQR = 4.8 × 10^8^-7.9 × 10^8^ cp/g) providing further support of efficient nucleic-acid extraction

### H5 RNA detection in wastewater samples

H5 RNA was first detected at the POTW on March 18, 2024 (**Fig 2**). Between March 18 and May 13 (the final sample of this study), 13 of 25 (52%) samples tested positive for H5 RNA. The median concentration detected was 2410 gc/dry g, with the peak concentration detected on May 3 (19,000 gc/dry g). H5 RNA detections were not related to rainfall patterns in the region (**Fig S1**).

**Figure 2.**
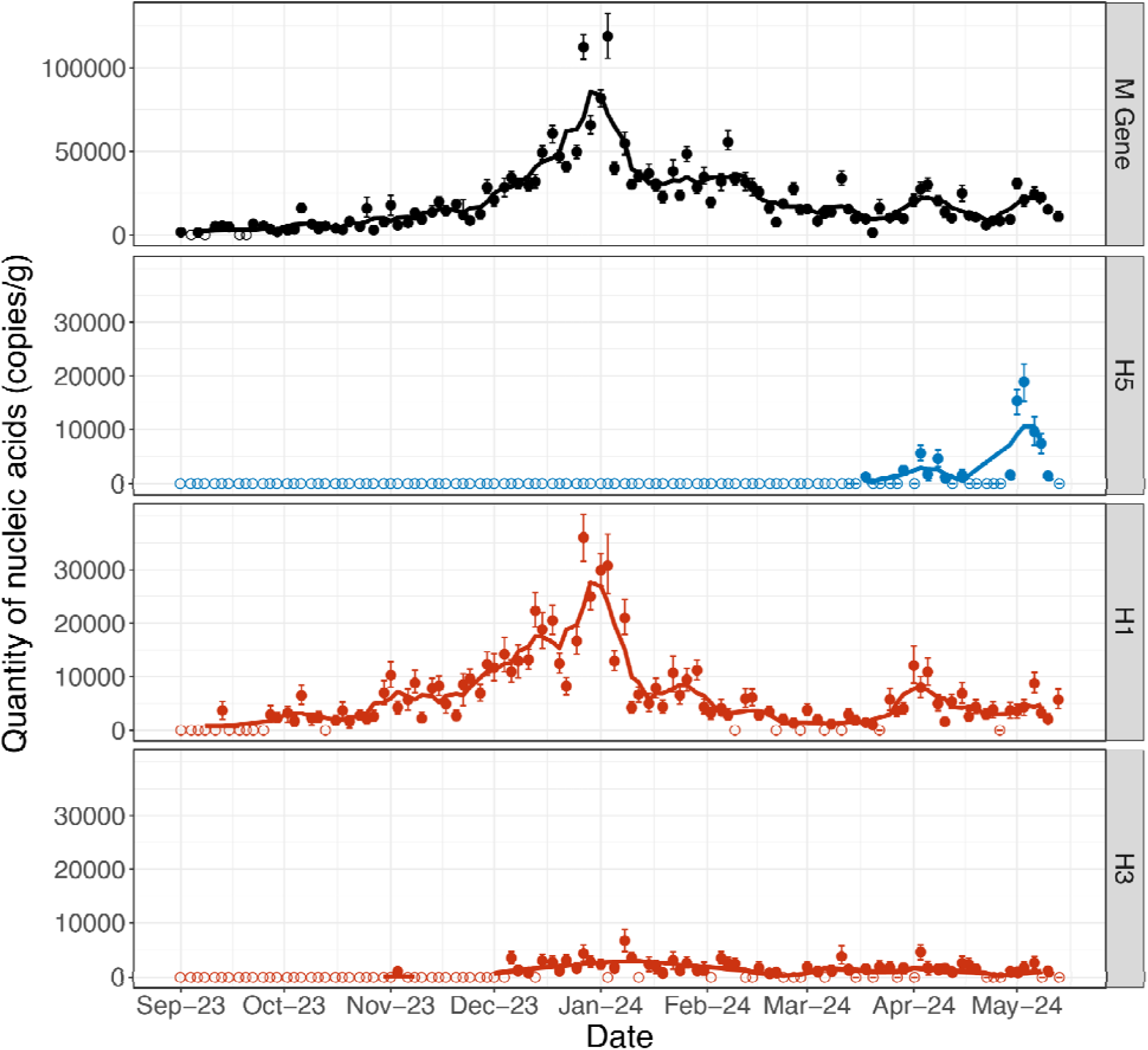
Concentrations of influenza A M gene (all IAV), H5, H1, and H3 RNA in wastewater solids. Concentrations measured are indicated by filled circles with the 68% confidence interval represented by bars. Samples with no detection are indicated by open circles. The lines represent the 5-adjacent sample trimmed average.

Both total IAV (M gene) and H1 RNA were regularly detected throughout the sampling period, and concentrations of both peaked in late December 2023 (**Fig 2**). H3 RNA was regularly detected from December 2023 onwards. H1 and H3 RNA concentrations remained low during H5 RNA detections from March through May. Of clinical influenza cases reported in CA during the 2023-2024 season, over 70% of cases were H1N1 (**Fig S2**)^42^.

### Modeled sources of H5 RNA

We calculated the total gene copies of H5 theoretically contributed to wastewater per infected human, liter of milk, and in daily fecal production in poultry and wild birds (**Table 3**). The estimated required inputs per source varied with measured H5 RNA concentrations (**Figure 3**). Overall, the lower bound (25th percentile) of contributions required to result in the median measured H5 RNA concentration were 10^4^ liters of milk, 10^4^ infected humans, 10^2^ infected poultry, or 10^5^ infected wild birds.

**Figure 3.**
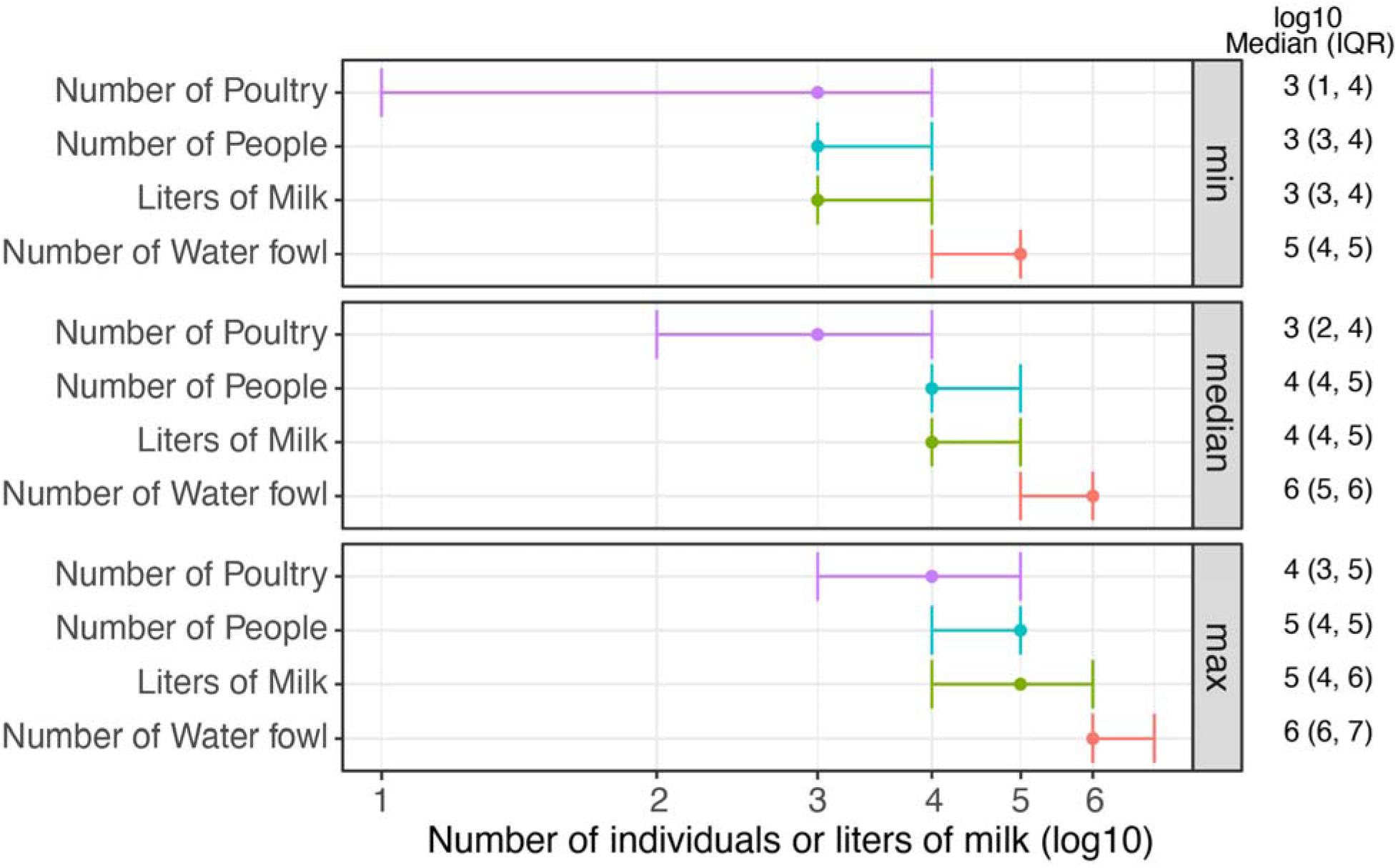
Median and IQR (25th - 75th quantile) of number of infected individuals required (humans, poultry, waterfowl) or liters of milk required to result in the lowest non-zero, median, and maximum H5 concentration measured at the POTW.

**Table 3.** Median gene copies of H5 RNA contributed to wastewater per day per infected individual (person, poultry, waterfowl) or per liter (IAV RNA containing milk).

| Source | Percentile |  |  |
| --- | --- | --- | --- |
|  | 25 | 50 (median) | 75 |
| Person | $2 \times 10^6$ | $9 \times 10^6$ | $3 \times 10^7$ |
| Liter of Milk | $7 \times 10^4$ | $4 \times 10^5$ | $2 \times 10^6$ |
| Poultry | $5 \times 10^6$ | $6 \times 10^7$ | $6 \times 10^8$ |
| Waterfowl | $9 \times 10^4$ | $3 \times 10^5$ | $1 \times 10^6$ |

The peak H5 RNA concentration was detected on May 3, 2024; for this concentration (19,000 gc/dry g), the lower bound (25th percentile) required contributions were 10^4^ liters of milk, 10^4^ infected humans, 10^3^ infected poultry, or 10^6^ infected waterfowl.

### Sensitivity Analysis

The H5 input model is very sensitive to the concentrations of IAV in poultry and waterfowl cloacal swabs, the concentration of influenza RNA in cow milk, and the concentration of influenza in human saliva (**Fig 4**).

**Figure 4.**
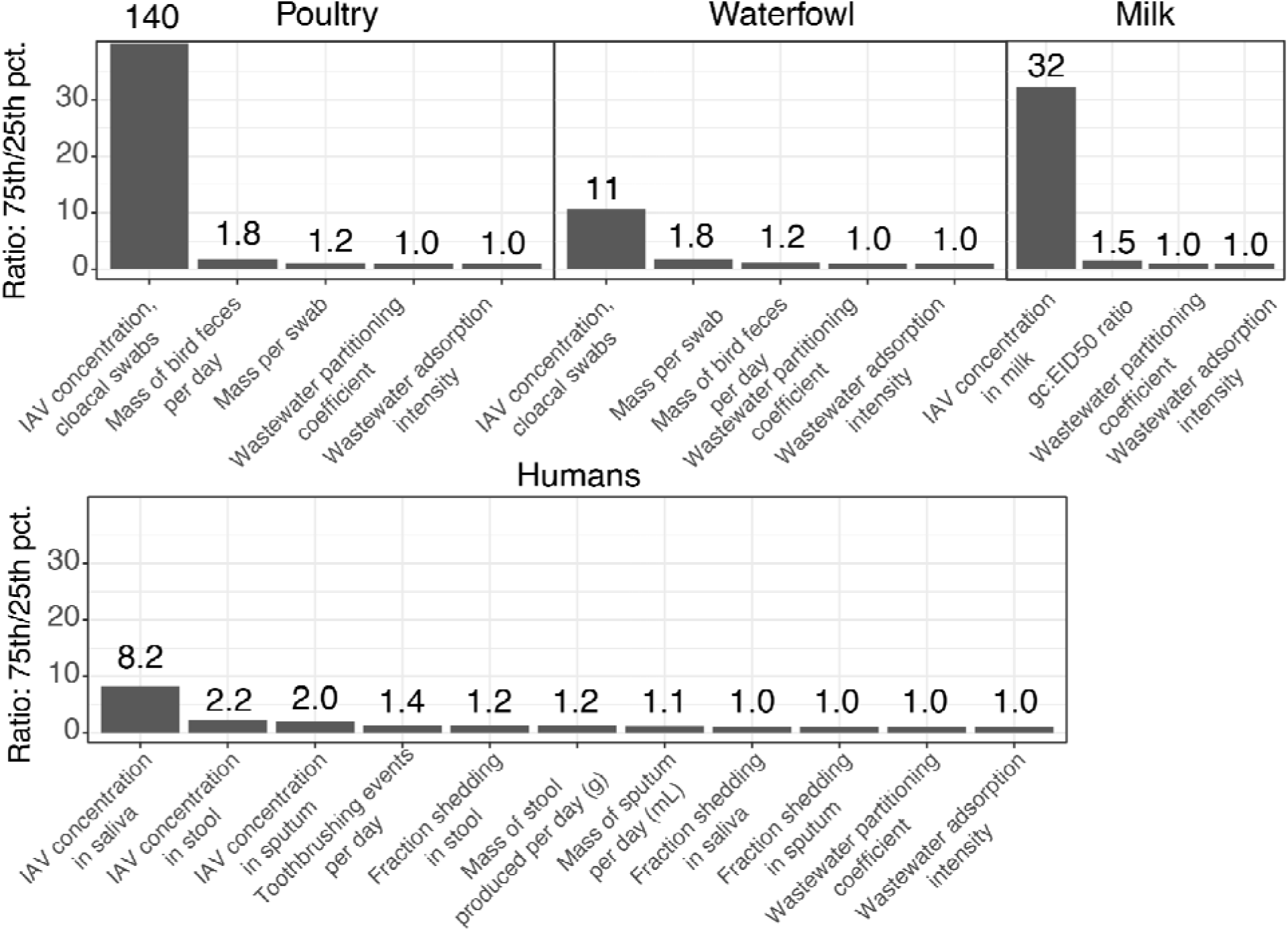
Sensitivity analysis results. Presented are the ratios of model output between the 75th and 25th percentiles for each parameter while all other parameters are set to their median value.

### Possible sources of animal inputs into the wastewater system

We identified two dairy processing facilities in the sewershed: a cheese making business and a butter creamery. We were not able to determine the volume of waste output from these businesses or whether their waste entered the sewer system.

Given that an estimated 20% of milk is wasted in residential settings,^19^ an estimated daily milk consumption per capita of 118 mL,^43^ and a sewershed population of 750,000, we estimate that approximately 22,000 liters of milk are wasted per day in households in the sewershed. This is a conservative estimate, not accounting for waste in pasteurization, dairy products, food service, or at retail stores (of which an estimated 15% is wasted^19^).

We also located one live bird market in the sewershed where poultry are kept alive and where poultry feces enter the sewershed via drains in the establishment. The market operates on select dates and birds are typically housed at the market for a maximum of 24 hours before being sold; thousands of birds can be present at any one time.

## Discussion

We first detected H5 RNA in San Francisco wastewater in mid-March 2024 after H5N1 outbreaks in cows had been identified in the United States but before any outbreaks had been reported in California. Samples from September 2023 to mid-March 2024 were all non-detect for H5 RNA, despite regular detection of the influenza A M gene and known outbreaks of H5N1 in domestic poultry and wild birds in the region.^8^ The timing of the initial H5 RNA detections generally aligned with detections of H5 RNA in wastewater from Amarillo, TX (with detections late February to late April in the South plant and detections from mid-March to late April in the North plant) and Dallas, TX (detections mid-March to mid-April).^16^ However, while dairy cow herds were identified with H5N1 illness in Texas during this time, none were identified in California until late August 2024.^44^ High concentrations of H5 RNA were detected in wastewater from dairy industries discharging to the wastewater system in Amarillo, TX,^16^ while no major dairy operations of similar scale were identified in the San Francisco sewershed. H5N1 outbreaks in poultry flocks were reported in San Francisco (May 9, 2024) within the time period of wastewater H5 RNA detections. As such, the source of H5 RNA in wastewater in San Francisco is unclear based on circumstantial evidence and we implemented a modeling approach to determine the feasibility of different H5 influenza sources to the sewer system producing these results. We found the model helpful to estimate the order of magnitude contributions that may be required from different pathways; however, data on shedding and other assumptions means that the ability of the model to make precise estimates is limited.

### Waterfowl

Based on the model results, waterfowl are unlikely to be a major source of H5 RNA to wastewater from the SF sewershed. Feces from at least ten thousand of these infected birds would be required to result in the minimum H5 RNA concentration measured, and there is unlikely to be a feasible way for the required mass of bird feces to enter the system despite it being a combined system in which some contribution is likely. This analysis was limited by a lack of data on wild bird fecal shedding of influenza. In California in 2024, waterfowl such as geese as well as owls, gull, falcons, and other types of wild birds have all tested positive for H5N1.^45^ Our shedding data was based on a study in blue-winged teals,^22^ which are considered representative of waterfowl.^31^ Influenza shedding in other types of birds with confirmed H5N1 in California are sparse.^23^ It is possible that other species of wildfowl could shed greater quantities of IAV in feces or produce much greater quantities of feces. However, the mechanisms through which wild bird feces enter the wastewater system are limited, and the model assumes all feces produced are entering the system. Stormwater or urban runoff entering the combined system could wash bird feces into the system, yet there were no precipitation events recorded in San Francisco during the period of H5 RNA detections.

Bird feces could enter the system through bodies of standing water that drain to the system, yet none were identified within the sewershed.

### Poultry

Poultry are a feasible contributor of H5 to the SF sewer system, especially given the live bird market located within the sewershed where chicken feces can be washed into the sanitary sewer system. Notably, poultry at the live bird market infected with H5N1 were identified on May 9, 2024,^8^ during the study period and coinciding with an H5 RNA wastewater detection. The USDA describes that the outbreak involved 700 birds;^8^ this number is within the IQR of the modeled number of poultry required for both the minimum and median H5 RNA concentrations measured in wastewater. Based on the author’s observation, we estimate that the bird market may house as many as 3,000 birds at one time, which is within the lower IQR (25th percentile) for the number of infected poultry required to result in the maximum H5 concentration measured.

However, there were additional H5 detections after the May 2024 outbreak and it is unlikely that such a large number of H5N1-infected birds could go undetected.

Outbreaks of highly pathogenic avian influenza (HPAI), which includes H5N1, result in severe morbidity and mortality in affected flocks. An early sign of HPAI is often unexpected chicken deaths, and common symptoms in chickens are severe and would be difficult to miss.^46^ On Dutch poultry farms, the time between virus introduction and final mortality within individual chicken flocks ranged from 5 - 12 days,^47^ signifying rapid spread through flocks. We saw sustained detection of H5 RNA in wastewater across 5 weeks in the POTW, which would require that chickens at the live bird market be consistently infected with H5N1 during this time period if the market were the sole or primary H5 RNA wastewater source.

The poultry model is very sensitive to poultry fecal shedding. Our model uses fecal shedding estimates for HPAI from a meta-analysis that incorporates data from 71 studies, including 25 studies specific to H5N1.^23^ The presentation of H5N1 in poultry in the U.S. since the beginning of the outbreak in 2022 has been similar to H5N1 outbreaks in poultry in other regions;^46^ as such, we feel that we are using the best available estimates of poultry fecal shedding.

### Milk

We find that milk is feasible as a primary or sole H5 RNA source to the POTW, but only if the milk supply is widely contaminated. Pasteurization of milk inactivates IAV, but does not reduce the IAV RNA concentration in milk which is still detectable by PCR.^48–50^ Residential milk waste could drive H5 RNA detections; even for the maximum H5 RNA wastewater concentration, the lower bound model output (25th percentile) requires 10,000 liters of contaminated milk to enter the sewer system. This is within our conservative estimate of the amount of milk wasted residentially each day in the sewershed (22,000 liters) not including the contribution of small dairies, retailers, and food service, and so it is possible that dumping of contaminated milk could result in the detected H5 RNA if there was widespread contamination of milk at retailers within SF.

This analysis was limited by the availability of data on H5 and IAV concentrations in milk. No H5 or IAV containing milk was reported in milk processed in California during the time of this study,^17,51^ although testing was limited (Tarbuck *et al*.: n=3, Suarez *et al*., n=6), and no infected dairy cows were reported in California during the time period of this study.^44^ However, it is possible that contaminated milk may enter the supply chain in San Francisco through either unidentified local cattle outbreaks or through milk processed in other locations with H5N1 outbreaks in dairy cattle. The model is sensitive to the H5 concentration in milk, and thus our input estimates would be affected by an over- or underestimation of the H5 concentration in milk.

### Humans

Infected humans are unlikely to be a major contributor of H5 RNA to the San Francisco wastewater. As of December 19, 2024, only 61 people in the United States have been identified with H5N1 infections, and these cases were mostly in people who worked closely with infected cows or poultry.^12^ Human-to-human transmission of H5N1 is not suspected at this time. While it is possible that asymptomatic or undiagnosed cases exist and the true number of infections is higher,^52^ there is no evidence to support the likelihood of widespread infection in humans at this time - especially 10^3^ cases as the model estimates would be required to produce the minimum H5 concentration.^13^

Our estimates of the required H5 human cases are constrained by available human IAV shedding data, and do not necessarily align with previous observations of influenza A in wastewater during outbreaks in humans. The shedding data are based on human shedding of influenza A overall (no subtyping conducted), H1N1, H3N2, or H7N9,^28^ and data on H5N1 shedding in humans are not available. No information is available on human IAV shedding of any subtype in urine.^28^ Recent H5N1 infections have presented differently than seasonally influenza,^13,53^ and shedding profiles of H5N1 may differ from those of other influenza types. We also noted that our estimate that 10^3^ to 10^4^ infected humans would be required to result in the minimum H5 concentration observed is 3-4 orders of magnitude larger than the number of infected individuals that were empirically observed to produce a similar concentration of total IAV in Ann Arbor, Michigan and at Stanford University, as determined through case reporting and in depth outbreak investigation.^2^ Future work is needed to better quantify human shedding of influenza in wastewater-relevant excretions.

Research is also needed into the possibility of human dietary shedding of H5 following consumption of H5N1-containing dairy products. Other single-stranded RNA viruses, such as PMMoV, brown rugose tomato virus, and porcine circoviruses, have been detected in human stool following consumption of foods containing these viruses.^54–57^ If H5 IAV present in milk or other dairy products is able to be shed in human stool, uninfected humans who consume dairy products could contribute to H5 wastewater detections.

## Limitations

We note that a major limitation of the study is the uncertainty in the data underlying many parameters, especially because the model was found to be sensitive to several of these related to shedding. These shedding parameters are particularly influential in the model, and in many cases are poorly understood. Concerns about this limitation are exacerbated by the observation that modeled estimates for human infections are substantially higher than would be expected based on observations from previous influenza A outbreaks of other subtypes. Despite these limitations, we feel that the model provides a helpful framework for discussing the order of magnitude estimated contributions from different sources to wastewater. More data on shedding in animals and humans will continue to improve estimates in the future.

Another limitation of this study is that inputs from humans, poultry, wild birds, and milk were estimated separately. Multiple sources could contribute to H5 RNA wastewater detections, yet there is no information available to constrain the fraction of input that may come from each source, and thus each source was modeled separately. If multiple sources contribute H5 to the wastewater system, the number of required infected individuals or liters of milk could be lower than the lower bounds estimated by our models. Our H5 assay will detect all IAV in the H5 subtype, and this could include strains other than the currently circulating H5N1 (e.g., H5N8); however, no other H5 strains are currently expected to be circulating in wastewater-relevant species. It should also be noted that the IAV genome, particularly the HA gene, has a high rate of mutation. Although the H1, H3, and H5 assays used herein are highly sensitive and specific for the dominate IAV H1, H3, and H5 subtypes circulating at the time of the study, they may not be highly sensitive and specific in different time periods during which different clades of each subtype are in circulation. For example, we have confirmed that the H1 assay is no longer sensitive for the H1N1 clade circulating in the 2024-2025 flu season.

The model developed could be applied to other sites with H5 detections in wastewater to help constrain potential sources, and we developed a Shiny app tool to allow the model to be easily deployed at other sites. For other sites using the model, however, the feasibility and appropriateness of each pathway included in the model would need to be determined based on the specifics of the wastewater system.

In this study, we show that H5 was detected in an urban California watershed months before H5N1 cases in California dairy cattle were reported. Given a lack of dairy industry in the sewershed or surrounding area, the source of H5 RNA in the wastewater system was unclear. Here, we employ modeling to estimate the theoretical H5 contributions from infected humans, poultry, and wildfowl, and from H5-contaminated milk. We demonstrate that humans and waterfowl are unlikely to be major contributors of H5 to the system, while poultry and milk are both feasible sources in this system.

## Data Availability

All data produced in the present study and EMMI guideline checklists are available online at https://purl.stanford.edu/ks454rq5640.

## Acknowledgements

We thank the Southeast San Francisco POTW staff for providing us with samples and the flow and total suspended solids data. We gratefully acknowledge all data contributors, i.e., the authors and their originating laboratories responsible for obtaining the specimens and their submitting laboratories for generating the genetic sequence and metadata and sharing via the GISAID Initiative, on which this research is based. This work was supported by a gift from the Sergey Brin Family Foundation to ABB. The graphical abstract and Figure 1 were created using Biorender.com.

## References

1. Tokars, J. I., Olsen, S. J. & Reed, C. Seasonal Incidence of Symptomatic Influenza in the United States. Clin. Infect. Dis. 66, 1511–1518 (2018).

2. Wolfe, M. K. et al. Wastewater-Based Detection of Two Influenza Outbreaks. Environ. Sci. Technol. Lett. 9, 687–692 (2022).

3. Toribio-Avedillo, D. et al. Monitoring influenza and respiratory syncytial virus in wastewater. Beyond COVID-19. *Sci. Total Environ.* **892**, 164495 (2023).

4. Lehto, K.-M. et al. Wastewater-based surveillance is an efficient monitoring tool for tracking influenza A in the community. Water Res. 257, 121650 (2024).

5. Maida, C. M. et al. Detection of influenza virus in urban wastewater during the season 2022/2023 in Sicily, Italy. Front. Public Health 12, (2024).

6. Nypaver, C., Dehlinger, C. & Carter, C. Influenza and Influenza Vaccine: A Review. J. Midwifery Womens Health 66, 45–53 (2021).

7. 7. CDC. Avian Influenza Type A Viruses. *Avian Influenza (Bird Flu)* https://www.cdc.gov/bird-flu/about/index.html (2024).

8. Confirmations of Highly Pathogenic Avian Influenza in Commercial and Backyard Flocks | Animal and Plant Health Inspection Service. https://www.aphis.usda.gov/livestock-poultry-disease/avian/avian-influenza/hpai-detections/commercial-backyard-flocks.

9. Lai, S. et al. Global epidemiology of avian influenza A H5N1 virus infection in humans, 1997–2015: a systematic review of individual case data. Lancet Infect. Dis. 16, e108–e118 (2016).

10. Monto, A. S. & Fukuda, K. Lessons From Influenza Pandemics of the Last 100 Years. Clin. Infect. Dis. 70, 951–957 (2020).

11. Peiris, J. S. M., de Jong, M. D. & Guan, Y. Avian Influenza Virus (H5N1): a Threat to Human Health. Clin. Microbiol. Rev. 20, 243–267 (2007).

12. 12. CDC. H5 Bird Flu: Current Situation. *Avian Influenza (Bird Flu)* https://www.cdc.gov/bird-flu/situation-summary/index.html (2024).

13. Garg, S. Outbreak of Highly Pathogenic Avian Influenza A(H5N1) Viruses in U.S. Dairy Cattle and Detection of Two Human Cases — United States, 2024. MMWR Morb. Mortal. Wkly. Rep. 73, (2024).

14. 14. CDC Newsroom. *CDC* https://www.cdc.gov/media/releases/2024/p-0703-4th-human-case-h5.html (2016).

15. 15. CDC Newsroom. *CDC* https://www.cdc.gov/media/releases/2024/p-0715-confirm-h5.html (2016).

16. Wolfe, M. K. et al. Detection of Hemagglutinin H5 Influenza A Virus Sequence in Municipal Wastewater Solids at Wastewater Treatment Plants with Increases in Influenza A in Spring, 2024. Environ. Sci. Technol. Lett. 11, 526–532 (2024).

17. 17. Tarbuck, N., et al. Detection of A(H5N1) influenza virus nucleic acid in retail pasteurized milk. Preprint at 10.21203/rs.3.rs-4572362/v1 (2024).

18. Spackman, E. et al. Characterization of highly pathogenic avian influenza virus in retail dairy products in the US. J. Virol. 98, e00881–24 (2024).

19. Thoma, G. et al. Greenhouse gas emissions from milk production and consumption in the United States: A cradle-to-grave life cycle assessment circa 2008. Int. Dairy J. 31, S3–S14 (2013).

20. Byker, C. J., Farris, A. R., Marcenelle, M., Davis, G. C. & Serrano, E. L. Food Waste in a School Nutrition Program After Implementation of New Lunch Program Guidelines. J. Nutr. Educ. Behav. 46, 406–411 (2014).

21. Shanks, C. B., Bark, K., Stenberg, M., Gamble, J. & Parks, C. Milk Consumption and Waste Across 5 Montana High School Lunch Programs. J. Sch. Health 90, 718–723 (2020).

22. Dolinski, A. C., Jankowski, M. D., Fair, J. M. & Owen, J. C. The association between SAα2,3Gal occurrence frequency and avian influenza viral load in mallards (Anas platyrhynchos) and blue-winged teals (Spatula discors). BMC Vet. Res. 16, 430 (2020).

23. Germeraad, E. A. et al. Virus Shedding of Avian Influenza in Poultry: A Systematic Review and Meta-Analysis. Viruses 11, 812 (2019).

24. Boehm, A. B. et al. Human viral nucleic acids concentrations in wastewater solids from Central and Coastal California USA. Sci. Data 10, 396 (2023).

25. Topol, A. High Throughput RNA Extraction and PCR Inhibitor Removal of Settled Solids for Wastewater Surveillance of SARS-CoV-2 RNA V2. https://www.protocols.click/view/high-throughput-rna-extraction-and-pcr-inhibitor-r-b2mkqc4w (2021) doi:10.17504/protocols.io.b2mkqc4w.

26. Boehm, A. B. et al. Human pathogen nucleic acids in wastewater solids from 191 wastewater treatment plants in the United States. Sci. Data 11, 1141 (2024).

27. Roldan-Hernandez, L., Oost, C. V. & B. Boehm, A. Solid–liquid partitioning of dengue, West Nile, Zika, hepatitis A, influenza A, and SARS-CoV-2 viruses in wastewater from across the USA. Environ. Sci. Water Res. Technol. (2024) doi:10.1039/D4EW00225C.

28. Lowry, S. A., Wolfe, M. K. & Boehm, A. B. Respiratory virus concentrations in human excretions that contribute to wastewater: a systematic review and meta-analysis. J. Water Health 21, 831–848 (2023).

29. Crank, K., Chen, W., Bivins, A., Lowry, S. & Bibby, K. Contribution of SARS-CoV-2 RNA shedding routes to RNA loads in wastewater. Sci. Total Environ. 806, 150376 (2022).

30. Dovas, C. I. et al. Detection and Quantification of Infectious Avian Influenza A (H5N1) Virus in Environmental Water by Using Real-Time Reverse Transcription-PCR. Appl. Environ. Microbiol. 76, 2165–2174 (2010).

31. Humphreys, J. M. et al. Waterfowl occurrence and residence time as indicators of H5 and H7 avian influenza in North American Poultry. Sci. Rep. 10, 2592 (2020).

32. Arena, C. et al. Simultaneous investigation of influenza and enteric viruses in the stools of adult patients consulting in general practice for acute diarrhea. Virol. J. 9, 116 (2012).

33. Hirose, R. et al. Long-term detection of seasonal influenza RNA in faeces and intestine. Clin. Microbiol. Infect. 22, 813.e1–813.e7 (2016).

34. Chan, M. C. W., Lee, N., Chan, P. K. S., Leung, T. F. & Sung, J. J. Y. Fecal detection of influenza A virus in patients with concurrent respiratory and gastrointestinal symptoms. J. Clin. Virol. 45, 208–211 (2009).

35. Chan, M. C. W. et al. Seasonal Influenza A Virus in Feces of Hospitalized Adults. Emerg. Infect. Dis. 17, 2038–2042 (2011).

36. Tańczuk, M., Junga, R., Kolasa-Wiecek, A. & Niemiec, P. Assessment of the Energy Potential of Chicken Manure in Poland. Energies 12, 1244 (2019).

37. Owen, R. B., Jr. The Bioenergetics of Captive Blue-Winged Teal under Controlled and Outdoor Conditions. The Condor 72, 153–163 (1970).

38. Julian, T. R., Canales, R. A., Leckie, J. O. & Boehm, A. B. A model of exposure to rotavirus from nondietary ingestion iterated by simulated intermittent contacts. Risk Anal. Off. Publ. Soc. Risk Anal. 29, 617–632 (2009).

39. Xue, J. et al. A Probabilistic Arsenic Exposure Assessment for Children Who Contact Chromated Copper Arsenate (CCA)-Treated Playsets and Decks, Part 2: Sensitivity and Uncertainty Analyses. Risk Anal. 26, 533–541 (2006).

40. Borchardt, M. A. et al. The Environmental Microbiology Minimum Information (EMMI) Guidelines: qPCR and dPCR Quality and Reporting for Environmental Microbiology. Environ. Sci. Technol. 55, 10210–10223 (2021).

41. McClary-Gutierrez, J. S. et al. Standardizing data reporting in the research community to enhance the utility of open data for SARS-CoV-2 wastewater surveillance. Environ. Sci. Water Res. Technol. 7, 1545–1551 (2021).

42. Health, D. of P. California Department of Public Health. https://www.cdph.ca.gov/Programs/CID/DCDC.

43. Stewart, H., Kuchler, F. & Hahn, W. Is competition among soft drinks, juices, and other major beverage categories responsible for reducing Americans’ milk consumption? Agribusiness 37, 731–748 (2021).

44. HPAI Confirmed Cases in Livestock | Animal and Plant Health Inspection Service. https://www.aphis.usda.gov/livestock-poultry-disease/avian/avian-influenza/hpai-detections/hpai-confirmed-cases-livestock.

45. HPAI Detections in Wild Birds. https://www.aphis.usda.gov/livestock-poultry-disease/avian/avian-influenza/hpai-detections/wild-birds.

46. Bordes, L. et al. Experimental infection of chickens, Pekin ducks, Eurasian wigeons and Barnacle geese with two recent highly pathogenic avian influenza H5N1 clade 2.3.4.4b viruses. Emerg. Microbes Infect. 13, 2399970 (2024).

47. Hobbelen, P. H. F. et al. Estimating the introduction time of highly pathogenic avian influenza into poultry flocks. Sci. Rep. 10, 12388 (2020).

48. Cui, P. et al. Does pasteurization inactivate bird flu virus in milk? Emerg. Microbes Infect. 13, 2364732 (2024).

49. Schafers, J. et al. Pasteurisation temperatures effectively inactivate influenza A viruses in milk. 2024.05.30.24308212 Preprint at 10.1101/2024.05.30.24308212 (2024).

50. Nooruzzaman, M. et al. Thermal inactivation spectrum of influenza A H5N1 virus in raw milk. 2024.09.21.614205 Preprint at 10.1101/2024.09.21.614205 (2024).

51. Suarez, D. L. et al. Testing of retail cheese, butter, ice cream and other dairy products for highly pathogenic avian influenza in the US. 2024.08.11.24311811 Preprint at 10.1101/2024.08.11.24311811 (2024).

52. Mellis, A. M. Serologic Evidence of Recent Infection with Highly Pathogenic Avian Influenza A(H5) Virus Among Dairy Workers — Michigan and Colorado, June–August 2024. MMWR Morb. Mortal. Wkly. Rep. 73, (2024).

53. Uyeki, T. M. et al. Highly Pathogenic Avian Influenza A(H5N1) Virus Infection in a Dairy Farm Worker. N. Engl. J. Med. 390, 2028–2029 (2024).

54. Zhang, T. et al. RNA Viral Community in Human Feces: Prevalence of Plant Pathogenic Viruses. PLOS Biol. 4, e3 (2005).

55. Colson, P. et al. Pepper Mild Mottle Virus, a Plant Virus Associated with Specific Immune Responses, Fever, Abdominal Pains, and Pruritus in Humans. PLOS ONE 5, e10041 (2010).

56. Li, L. et al. Multiple Diverse Circoviruses Infect Farm Animals and Are Commonly Found in Human and Chimpanzee Feces. J. Virol. 84, 1674–1682 (2010).

57. Natarajan, A. et al. The Tomato Brown Rugose Fruit Virus Movement Protein Gene Is a Novel Microbial Source Tracking Marker. Appl. Environ. Microbiol. 89, e00583–23 (2023).

