## Supplementary Information for "Detection of Hemagglutinin H5 influenza A virus RNA and model of potential inputs in an urban California sewershed"

***ddPCR Reaction Chemistry and Cycling Parameters***

The IAV M gene probe contained the fluorescent molecule ROX, H1 contained ATTO590, H3 contained both FAM and HEX, and H5 contained FAM; all probes also contained ZEN, an internal quencher from IDT, and IBFQ, Iowa Black FQ. Digital droplet RT-PCR methods have been described in detail elsewhere.^1^ Briefly, 20 μl of a 22 μl reaction volume are used for ddPCR; the reaction consists of 5.5 μl template, 5.5 μl of One-Step RT-ddPCR Advanced kit for Probes, 2.2 μl reverse transcriptase, 1.1 μl dithiothreitol, and primers and probes at a final concentration of 900 nM and 250 nM, respectively. The AutoDG Automated Droplet Generator was used to generate droplets. PCR was performed using Mastercycler Pro with the following protocol: reverse transcription at 50 °C for 60 min, enzyme activation at 95 °C for 5 min, 40 cycles with 1 cycle consisting of denaturation at 95 °C for 30 s and annealing and extension at either 59 °C for 30 s, enzyme deactivation at 98 °C for 10 min, and then an indefinite hold at 4 °C. The ramp rate for temperature changes was set at 2 °C/s, and the final hold at 4 °C was performed for a minimum of 30 min to allow the droplets to stabilize. Droplets were analyzed using either the QX200 or QX600 Droplet Reader (Bio-Rad). All liquid transfers were performed using the Agilent Bravo (Agilent Technologies).

***Additional details related to EMMI guidelines***

The average (standard deviation) number of droplets across the 10 replicate wells was 165,961 (26,274) for the multiplex reaction. The volume of each droplet is 0.00085 μL, as reported by the manufacturer (BioRad). The average (standard deviation) copies per droplet was 9.3x10^-8^ (3.8x10^-7^).

***Precipitation Data***

The daily precipitation dataset for San Francisco was obtained from the NOAA National Centers for Environmental Information Climate Data Online tool for the San Francisco Downtown, CA US site (ID: USW00023272).^2^

***Clinical Data***

Clinical cases by MMWR week for the 2023-2024 season were obtained for the state of California from the California Department of Public Health Influenza Surveillance Program.^3^

***Fitting Distributions to Parameters***

*Extracting raw data*

We obtained raw data from sources whenever possible. When raw data were not available in publications but results were plotted in figures, we used the WebPlotDigitizer tool to extract values from figures.^4^

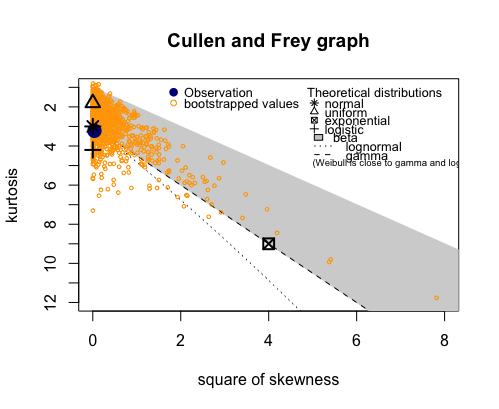

**Figure S1.** Example Cullen and Fray graph produced by the *descdist* function in the *fitdistrplus* package in R.

*Parameters with limited or no data available*

Influenza shedding in human urine

We could find no information on the quantities of influenza shed via urine for humans. We identified one study that reported shedding in human urine, yet values were not presented in externally valid units and as such could not be used for these estimations.^5^ Of note, the authors report quantities of IAV in urine that are 3+ orders of magnitude lower than in nasal or throat swabs. We choose to make the simplifying assumption that shedding in urine is negligible and set the shedding parameter equal to zero.

Mass of feces per cloacal swab, ${CF}_{SG}$

We can find no data on the mass of feces collected in a poultry cloacal swab. Therefore, we base the value of this parameter on our own lab experience and assume a uniform distribution ranging from 0.05 to 0.2 grams of feces per swab.

Daily volume of poultry fecal production, $V_{f}$

Volumes of feces produced per chicken per day are estimated from Table 2 of Tanczuk *et al*.^6^ Tanczuk et al. report a range of daily fecal volume per bird depending on bird type, with laying hens producing 150g per day and meat chicken producing 160g per day. Variability in fecal production by bird type is not reported. Authors assume that addition of straw litter in poultry areas increases fecal production by 15%. We assume that the range of fecal production varies 15% below the minimum produced amount present in Tanczuk et al. and 15% above the maximum. Therefore, we assume a uniform concentration ranging from 130 to 184g of daily fecal volume per bird.

Daily volume of waterfowl fecal production, $V_{f}$

Estimates of waterfowl fecal production are based on Owen, 1970, which examined energy production of the blue-winged teal.^7^ Blue-winged teals are considered representative of waterfowl.^8^ Owen measured the mass of feces produced in 3 days by the blue-winged teal across a range of temperatures (10°C, 20°C, and 30°C). We extracted raw data on fecal production / 3 days by temperature using WebPlotDigitizer. From these data, we assume a uniform distribution ranging from the minimum production reported (at 30°C) to the maximum production reported (at 10°C).

Fractional shedding in poultry and waterfowl

Given the lack of available data on the fraction of infected poultry or waterfowl shedding influenza in feces, we make the simplifying assumption that all infected birds shed in feces.

Concentration of influenza in poultry feces, $C_{CS}$

We used the values presented in a meta-analysis by Germeraad et al.^9^ Values were extracted from Figure 3 for chicken shedding via cloacal swabs for HPAI; boxplots presented the median and 25^th^ and 75^th^ percentile of gc/swab in poultry cloacal samples. The data presented appeared normal and so we assumed the data followed a normal distribution. Using WebPlotDigitizer, we extracted values for the mean/median, 25^th^ percentile, and 75^th^ percentile. We calculated the standard deviation of the dataset by solving for sd in the following equation: $mean+0.67\times sd= 75th percentile$. Data were fit to a normal distribution based on the mean and calculated sd.

Concentration of influenza in blue-winged teal cloacal swabs, $C_{CS}$

Concentrations of influenza in blue-winged teal cloacal swabs were estimated from Additional File 2 of Dolinski *et al*.^10^ The mean and upper and lower 95% confidence intervals were extracted from the plot using WebPlotDigitizer, and the total number of birds included in the experiment, *n*, was extracted from the paper’s methods. The standard deviation of the data was calculated by:

$$mean\pm t_{95\%}\times\frac{sd}{\sqrt{n}}={CI}_{95}$$

where ${CI}_{95}$ is the extracted values for the 95% confidence interval and is the standard deviation. By rearranging,

$$sd=\frac{({CI}_{95}\pm mean)\times\sqrt{n}}{t_{95\%}}$$

By plugging in both the upper and lower values, we calculate two values for sd: 0.72 (using upper estimate) and 0.80 (using lower estimate). We take the average of these two values to arrive at an estimate of sd = 0.76. Data are assumed to follow a normal distribution and the mean and calculated sd are used for the distribution.

Partitioning coefficient, $K_{f}$, and intensity of adsorption, *n*

Freundlich partitioning coefficients were retrieved from Table S5 of Roldan-Hernandez *et al*.^11^ Values for the intensity of adsorption (*n*) were retrieved from Table S6 of Roldan-Hernandez *et al*. Values were assumed to follow a uniform distribution with the minimum and maximum values equivalent to the minimum and maximum reported.

**Table S1.** Primers and probes for the H1, H3, and H5 triplex. The amplicon lengths are 106 bp for M gene, 178 bp for H1, 171 bp for H3, and 168 bp for H5.

| M Gene | Forward | CAAGACCAATCYTGTCACCTCTGAC |
| --- | --- | --- |
|  | Reverse | GCATTYTGGACAAAVCGTCTACG |
|  | Probe | TGCAGTCCTCGCTCACTGGGCACG |
| H1 | Forward | GTGAATCACTCTCCACAGCA |
|  | Reverse | TGATTRGGCCATGAACTTGT |
|  | Probe | TGGAACKTGTTACCCAGGAGA |
| H3 | Forward | GAGATCAGAYGCACCCATTG |
|  | Reverse | TCWGGTACATTYCGCATCCC |
|  | Probe | TGCATCACTCCAAATGGAAGCA |
| H5 | Forward | TATAGARGGAGGATGGCAGG |
|  | Reverse | ACDGCCTCAAAYTGAGTGTT |
|  | Probe | AGGGGAGTGGKTACGCTGCRGAC |

**Table S2.** Panel of pathogens used for in vitro specificity testing. All are inactivated pathogens from Zeptometrix (Buffalo, NY) unless “Twist” appears in front of the name in which case it is nucleic acid purchased from Twist (South San Francisco, CA).

| Parainfluenza 1  Parainfluenza 2  Parainfluenza 3  Parainfluenza 4  Influenza A H1N1  Influenza A H1  Influenza A H3  Influenza B  Adenovirus 1  Adenovirus 3 | Adenovirus 31  Rhinovirus Type 1A  RSV A  RSV B  SARS-CoV-2  *M. pneumoniae*  *C. pneumoniae*  Metapneumovirus 8  Coronavirus HKU-1  Coronavirus 229E | Coronavirus NL63  Coronavirus OC43  *B. parapertussis*  *B. pertussis*  Twist Influenza B  Twist Influenza A H1N1  Twist Influenza A H3N2 |
| --- | --- | --- |

**Table S3.** Positive control material

| Assay | Source/sequence |
| --- | --- |
| M Gene | Twist Synthetic Influenza H3N2 RNA Control (Twist 103002) |
| H5 | Custom gblock*  Set 1: AGAGAGGACTATTTGGAGCTATAGCAGGTTTTATAGAGGGAGGATGGCAGGGAATGGTAGATGGTTGGTATGGGTACCACCATAGCAATGAGCAGGGGAGTGGGTACGCTGCAGACAAAGAATCCACTCAAAAGGCAATAGATGGAGTCACCAATAAGGTCAACTCGATCATTGACAAAATGAACACTCAGTTTGAGGCCGTTGGAAGGGAATTTAATAACTTAGAAAGGAGAATAGAGAATT  Set 2:  AGAGAGGACTATTTGGAGCTATAGCAGGTTTTATAGAGGGAGGATGGCAGGGAATGGTAGATGGTTGGTATGGGTACCACCATAGCAATGAGCAGGGGAGTGGGTACGCTGCAGACAAAGAATCCACTCAAAAGGCAATAGATGGAGTCACCAATAAGGTCAACTCGATCATTGACAAAATGAACACTCAATTTGAGGCCGTTGGAAGGGAATTTAATAACTTAGAAAGGAGAATAGAGAATT |
| H1 | Twist Synthetic Influenza H1N1 RNA Control (Twist 103001) |
| H3 | Twist Synthetic Influenza H3N2 RNA Control (Twist 103002) |

* Two positive controls were generated and used in equimolar concentrations to account for a degeneracy present in the primer/probe design.

**Table S5.** Relevant SIC Codes for NPDES Permits

| **SIC** | **Description** |
| --- | --- |
| 0211 | Beef cattle feedlots |
| 0212 | Beef cattle, except feedlots |
| 0241 | Dairy farms |
| 2011 | Meat packing plants |
| 2013 | Sausages and other prepared meats |
| 2015 | Poultry slaughtering and processing |
| 2021 | Creamery butter |
| 2022 | Cheese, natural and processed |
| 2023 | Dry, condensed, evaporated products |
| 2024 | Ice cream and frozen desserts |
| 2026 | Fluid milk |
| 2077 | Animal and marine fats and oiles |
| 2079 | Edible fasts and oils, nec |
| 0213 | Hogs |
| 0214 | Sheep and goats |
| 0219 | General livestoc, nec |
| 0251 | Boiler, fryer, and roaster chickens |
| 0252 | Chicken eggs |
| 0253 | Turkeys and turkey eggs |
| 0254 | Poultry hatcheries |
| 0259 | Poultry and eggs, nec |
| 0271 | Fur-bearing animals and rabbits |
| 0272 | Horses and other equines |
| 0273 | Animal aquaculture |
| 0279 | Animal specialties, nec |
| 0291 | General farms, primarily animal |
| 0741 | Veterinary services for livestock |
| 0742 | Veterinary services, specialties |
| 0751 | Livestock services, exc. Veterinary |
| 0752 | Animal specialty services |
| 5143 | Dairy products, exc. Dried or canned, wholesale trade |
| 5144 | Poultry and poultry products, wholesale trade |
| 5147 | Meats and meat products, wholesale trade |
| 5154 | Livestock, wholesale trade |
| 5421 | Meat and fish markets |
| 5451 | Dairy product stores |

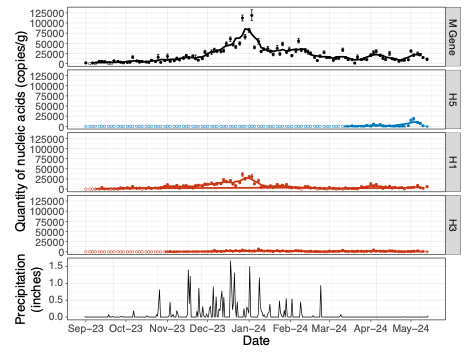

**Figure S1.** Concentrations of IAV: total IAV (M gene), H5, H1, and H3, and daily precipitation (inches) in San Francisco.

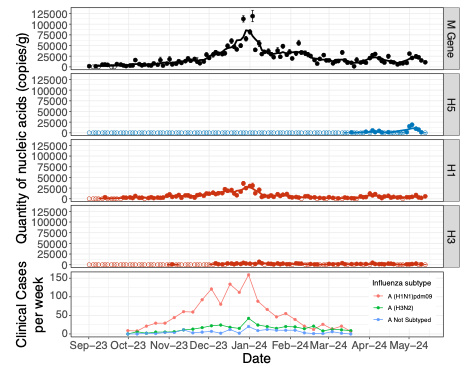

**Figure S2.** Concentrations of IAV: total IAV (M gene), H5, H1, and H3, and weekly clinical cases of type A Influenza in California. Data for clinical cases were available for October 1, 2023 – March 17, 2024.

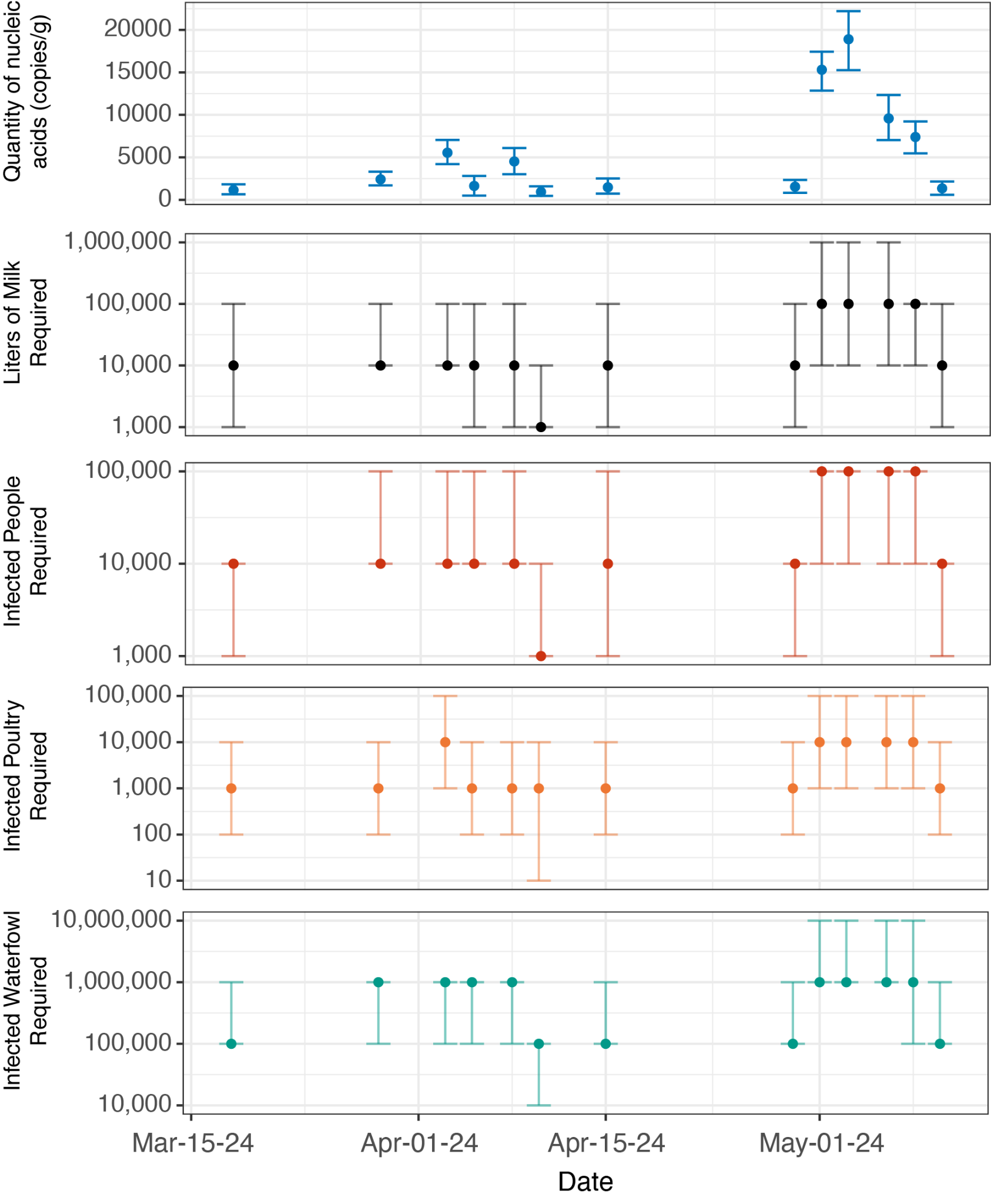

**Figure S3.** Source model results over time. Error bars on panels of model results (panels 2 - 5) represent IQR (25th - 75th percentile), while error bars on the H5 concentration panel (top) represent 95% confidence intervals around the measured concentration.

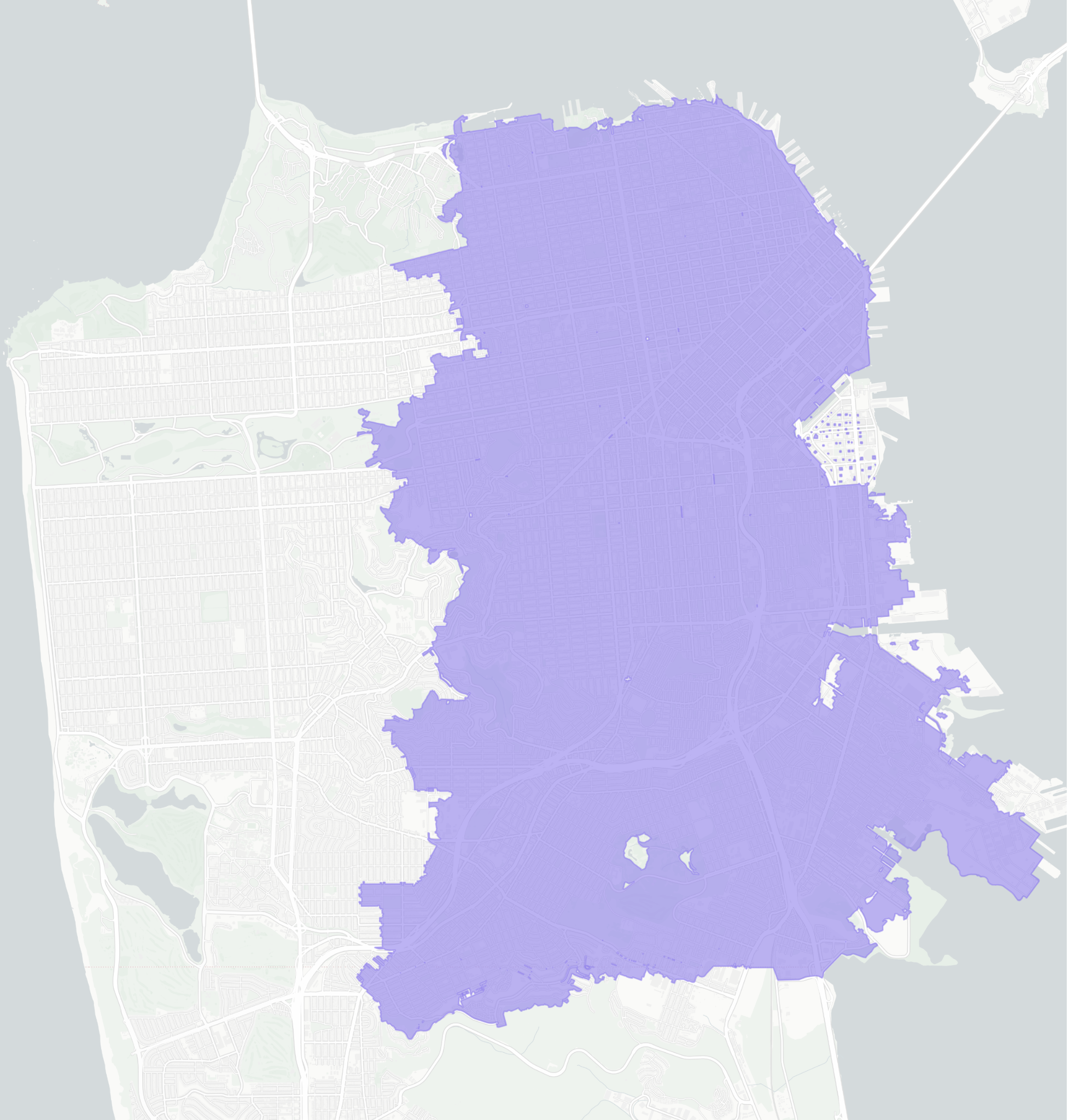

**Figure S4.** Map of the southeast San Francisco sewershed (data.wastewaterscan.org).

**References**

1.	Boehm, A. B. *et al.* Human pathogen nucleic acids in wastewater solids from 191 wastewater treatment plants in the United States. *Sci. Data* **11**, 1141 (2024).

2.	NOAA NWS. National Weather Service. *Climate Services* https://w2.weather.gov/climate/.

3.	Health, D. of P. California Department of Public Health. https://www.cdph.ca.gov/Programs/CID/DCDC.

4.	Rohatgi, A. WebPlotDigitizer.

5.	Kumar, B. *et al.* Quantification of viral load in clinical specimens collected from different body sites of patients infected with influenza viruses. *Int. J. Med. Med. Sci.* **3**, 144–148 (2011).

6.	Tańczuk, M., Junga, R., Kolasa-Wiecek, A. & Niemiec, P. Assessment of the Energy Potential of Chicken Manure in Poland. *Energies* **12**, 1244 (2019).

7.	Owen, R. B., Jr. The Bioenergetics of Captive Blue-Winged Teal under Controlled and Outdoor Conditions. *The Condor* **72**, 153–163 (1970).

8.	Humphreys, J. M. *et al.* Waterfowl occurrence and residence time as indicators of H5 and H7 avian influenza in North American Poultry. *Sci. Rep.* **10**, 2592 (2020).

9.	Germeraad, E. A. *et al.* Virus Shedding of Avian Influenza in Poultry: A Systematic Review and Meta-Analysis. *Viruses* **11**, 812 (2019).

10.	Dolinski, A. C., Jankowski, M. D., Fair, J. M. & Owen, J. C. The association between SAα2,3Gal occurrence frequency and avian influenza viral load in mallards (Anas platyrhynchos) and blue-winged teals (Spatula discors). *BMC Vet. Res.* **16**, 430 (2020).

11.	Roldan-Hernandez, L., Oost, C. V. & B. Boehm, A. Solid–liquid partitioning of dengue, West Nile, Zika, hepatitis A, influenza A, and SARS-CoV-2 viruses in wastewater from across the USA. *Environ. Sci. Water Res. Technol.* (2024) doi:10.1039/D4EW00225C.
